# Effects of Benzodiazepine Exposure on Real-World Clinical Outcomes in Individuals at Clinical High-Risk for Psychosis

**DOI:** 10.1101/2023.08.15.23294108

**Authors:** Nicholas R. Livingston, Andrea De Micheli, Robert McCutcheon, Emma Butler, Marwa Hamdan, Anthony A. Grace, Philip McGuire, Alice Egerton, Paolo Fusar-Poli, Gemma Modinos

## Abstract

**Background:** Animal models indicate GABAergic dysfunction in the development of psychosis, and that benzodiazepine (BDZ) exposure can prevent the emergence of psychosis-relevant phenotypes. However, whether BDZ exposure influences the risk of psychosis in humans is unknown.

**Methods:** This observational-cohort study used electronic health record data from 818 individuals at clinical high-risk for psychosis (CHR-P) to investigate whether BDZ exposure (including hypnotics e.g., zopiclone) reduces the risk of developing psychosis and adverse clinical outcomes. Cox proportional-hazards models were employed in both the whole-unmatched sample, and a propensity score matched (PSM) subsample.

**Results:** 567 CHR-P individuals were included after data cleaning (105 BDZ-exposed, 462 BDZ-unexposed). 306 (54%) individuals were male, and the mean age was 22.3 years (SD 4.9). The BDZ-exposed and BDZ-unexposed groups differed on several demographic and clinical characteristics, including psychotic symptom severity. In the whole-unmatched sample, BDZ exposure was associated with increased risk of transition to psychosis (HR=1.61; 95%CI:1.03-2.52; *P*=0.037), psychiatric hospital admission (HR=1.93; 95%CI:1.13-3.29; *P*=0.017), home visit (HR=1.64; 95%CI:1.18-2.28; *P*=0.004), and A&E attendance (HR=1.88; 95%CI:1.31-2.72; *P*<0.001). However, after controlling for confounding-by-indication through PSM, BDZ exposure did not modulate the risk of any outcomes (all *P*>0.05). In analysis restricted to antipsychotic-naïve individuals, BDZ exposure *reduced* the risk of transition to psychosis at trend-level (HR=0.59; 95%CI:0.32-1.08; *P*=0.089).

**Conclusions:** BDZ exposure in CHR-P individuals was not associated with a reduction in the risk of psychosis transition or other adverse clinical outcomes. Results in the whole-unmatched sample suggest BDZ prescription may be more likely in CHR-P individuals with higher symptom severity.

## 1. Introduction

Psychotic disorders are amongst the most severe psychiatric disorders, associated with chronic functional disability^1^, poor physical health, reduced life expectancy^2^, and high personal and familial burden^3^. Efforts to prevent the onset of psychosis in those at clinical high-risk (CHR-P) have so far been unsuccessful^4,5^. CHR-P individuals display attenuated psychotic symptoms or a first-degree familial risk, and about 25% of them will develop psychosis within 3 years^6,7^. Almost 50% of those who do not develop psychosis remain in the CHR-P state^8^, associated with reduced functioning and quality of life^9^, neurocognitive impairments^10,11^, and increased mental health resource utilisation, including Accident and Emergency department (A&E) attendance and psychiatric hospital admission^12,13^. However, there is no evidence to favour any available interventions for reducing transition to psychosis^14,15^. There is thus a substantial need to develop interventions to prevent the onset of psychosis and improve clinical outcomes in CHR-P individuals.

Psychotic symptoms are associated with increased dopamine release in the striatum^16^, which is likely to occur from upstream pathophysiological mechanisms^17^. A key mechanism is dysfunction of the gamma-aminobutyric acid (GABA) system in the hippocampus^18^, leading to increased glutamatergic drive to the striatum, and consequently increased striatal dopaminergic neuron firing and dopamine release^19^. Preclinical work in a well-validated rodent neurodevelopmental model – involving embryonic administration of methylazoxymethanol acetate (MAM) – with relevance to psychosis has demonstrated that peripubertal repeated administration of the GABA-enhancing drug diazepam, a benzodiazepine (BDZ), can prevent the emergence of both dopamine system hyperresponsivity and psychosis-relevant behavioural phenotypes at adulthood^20–22^. Additionally, the therapeutic effect of a more specific compound targeting α5GABA_A_ receptor subunits in MAM-treated rats is blocked by previous exposure to the antipsychotic drug haloperidol^23^. These findings suggest that enhancing GABAergic signalling during the premorbid phase of psychosis, prior to antipsychotic treatment, may prevent the development of psychosis. Whether this therapeutic potential of BDZs at the preclinical level translates to humans remains to be investigated.

Based on the above preclinical findings^20,21,23,24^, we conducted a naturalistic, retrospective, observational cohort-design study using electronic health record (EHR) data from a large sample of CHR-P individuals to investigate the effect of BDZ exposure on clinical outcomes. We hypothesised that BDZ exposure would reduce the risk of transition to psychosis and events indicative of a clinical crisis: psychiatric hospital admission, home visit, and A&E attendance. Due to preclinical evidence suggesting a masking effect by prior antipsychotic treatment^23^, a sensitivity analysis was also performed excluding CHR-P individuals with prior antipsychotic exposure.

## 2. Methods

The authors assert that all procedures contributing to this work comply with the ethical standards of the relevant national and institutional committees on human experimentation and with the Helsinki Declaration of 1975, as revised in 2008. All procedures were approved by South London and Maudsley NHS Foundation Trust Psychosis CAG (PSYAUD17_25).

### 2.1 Study Design, Setting, and Population

This observational cohort study used EHR data from CHR-P individuals accessing OASIS (Outreach and Support in South London)^25^, a CHR-P service within the South London and Maudsley National Health Service Foundation Trust, London, UK^26^. Data collection was between 2001-2021, and final data analysis was completed September-December 2022. To allow for a minimum follow-up period of 12-months, individuals who joined OASIS after September 2021 were excluded. Details regarding data cleaning and a schematic timeline of the exposure/observation periods are presented in Figure 1.

**Figure 1.**
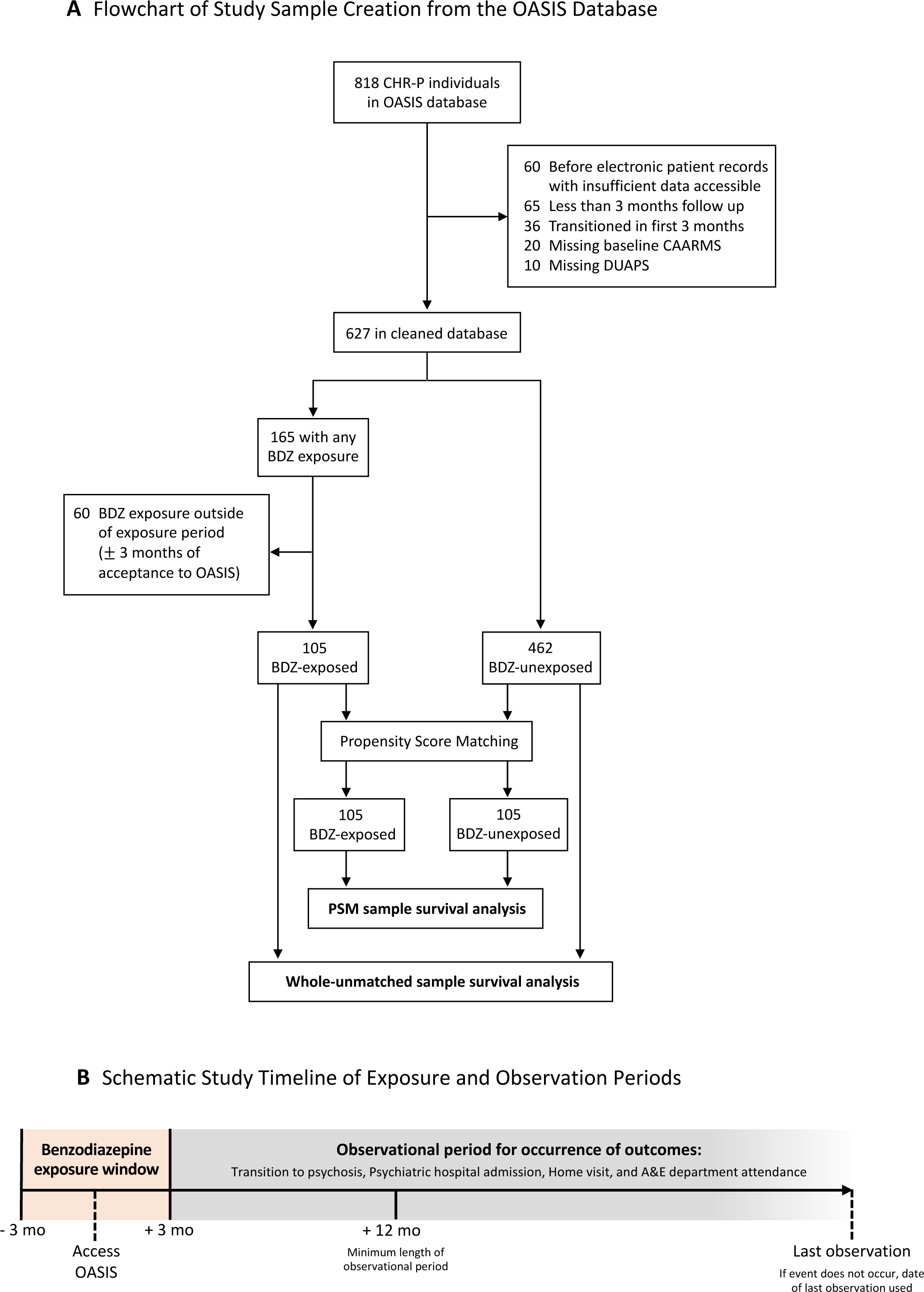
Study Design. Flowchart of Study Sample Creation from the OASIS Database (A) and Schematic Study Timeline of Exposure and Observation Periods (B). Reasons for exclusion of ineligible participants from the study and survival analyses completed in the whole-unmatched sample and the subset sample created through propensity score matching. Benzodiazepine exposure window was operationalised as ± 3-months of time joining OASIS. Observational periods begun at 3-months after accessing OASIS, and minimum follow up period was operationalised at 9-months after this. Time-to-event was used for primary and secondary outcomes, and when an event did not occur date of last observation was used. BDZ: Benzodiazepine; CAARMS: Comprehensive Assessment of At-Risk Mental State; CHR-P: Clinical High Risk for Psychosis; DUAPS: Duration of Untreated Attenuated Psychotic Symptoms; OASIS: Outreach And Support In South London; PSM: Propensity Score Matching

### 2.2 Benzodiazepine Exposure

BDZ exposure was operationalised as ≥ 1 dose of a BDZ within the exposure window (3-months prior to 3-months after accessing OASIS), and total number of days of BDZ exposure was calculated across the exposure window. BDZ exposure included non-benzodiazepine hypnotics e.g., zopiclone, due to their very similar pharmacological mechanism of action via full agonism of the benzodiazepine binding site on GABA_A_ receptors^27^. BDZ-unexposed individuals were defined as having no recorded exposure to BDZs in the time prior to accessing OASIS up until the end of their observation period (3-months after accessing OASIS to date of last event, or when an event did not occur, date of last observation). Individuals who received a BDZ outside the exposure window but within the observation period were removed from the analysis.

### 2.3 Primary and Secondary Outcomes

The primary outcome was diagnosis of an ICD-10 psychotic disorder within the observation period. Secondary outcomes were psychiatric hospital admission, home visit, and A&E attendance. A home visit is undertaken in instances when there is an indication of deterioration in the client’s mental state in the presence of risk.

### 2.4 Statistical Analysis

Statistical analyses were carried out using R (version 4.2.2). All statistical tests were two-sided and statistical difference was set at *P*<.05.

#### 2.4.1 Propensity Score Matching

As this study used real-world observational data, clinical and demographic factors may influence both the likelihood of BDZ exposure and the study’s primary and secondary outcomes, resulting in confounding-by-indication. To address this, a comparator BDZ-unexposed group was created through propensity score matching (PSM)^28^ using the MatchIt^29^ package in R (version 4.2.2). PSM involves running a logistic regression model on the whole database, with the binary outcome variable indicating BDZ exposure within the exposure window. The following covariates were entered into the model, which have been previously associated with BDZ prescription and/or our clinical outcome variables of interest^12^: attenuated psychotic symptom severity when joining OASIS (specifically the total score of ’unusual thought content’ and ‘non-bizarre ideas’ sub-scales^30,31^ on the CAARMS^32^ [Comprehensive Assessment of At Risk Mental States] instrument), age, black ethnicity, duration of untreated attenuated psychotic symptoms (DUAPS), date of joining OASIS, and the occurrence of brief limited intermittent psychotic symptoms (BLIPS). The model estimates were then used to calculate a propensity score for each individual, which represents the predicted probability of being exposed to a BDZ given these covariates. Each BDZ-exposed individual was then matched using the nearest neighbour method with a BDZ-unexposed individual with a near identical propensity score, resulting in two equally-sized matched groups. The success of the PSM was assessed as the similarity of covariates between the groups using chi-square and *t* tests for categorical and continuous variables, respectively.

#### 2.4.2 Survival Analysis

Cox proportional-hazards models investigated whether BDZ exposure modulated the risk of primary and secondary outcomes. Individual models were run for each outcome, in the whole-unmatched sample and in the PSM sample. The proportional-hazard assumption was used to check that hazards remained constant over time for each outcome.

#### 2.4.3 Sensitivity Analyses

In a primary sensitivity analysis, individuals with antipsychotic exposure prior to their BDZ exposure were removed from the database before statistical analysis. Three supplementary sensitivity analyses were run: i.) minimum 3 and ii.) 7 total days of BDZ exposure, and iii.) removing non-benzodiazepine hypnotics (e.g., zopiclone).

## 3. Results

### 3.1 Data cleaning and final sample characteristics

From the 818 CHR-P individuals in the OASIS database, 567 individuals were included (mean age 22.3 years [SD 4.9]; 306 [54%] male; 261 [46%] female) following data cleaning (Figure 1; Table 1). This included 105 individuals with BDZ exposure (mean follow up = 1157 [SD 1070] days) and 462 BDZ-unexposed individuals (mean follow up = 1264 [SD 1211] days). The median total number of days of BDZ exposure was 7 (IQR 3-21) days across the whole six-month exposure window. The most common BDZ was zopiclone (51%), and the most common reason for prescription was for sleep (56%; Supplementary Table 1). Within the final database of 567 individuals, compared to the BDZ-unexposed group, the BDZ-exposed individuals were more likely to be older (mean 24.3 vs. 21.9 years), of black ethnicity (46 [44%] vs. 148 [32%]), be classified within the BLIPS CHR-P subgroup (45 [43%] vs. 55 [12%]), have higher psychotic symptom severity at baseline assessment (mean 9.2 vs. 7.6 score out of 12), have a shorter DUAPS (mean 392 vs. 338 days), and have accessed OASIS more recently (mean 42345 vs. 41964 timepoint, made by converting date to numerical number). After PSM, there was no statistical difference between the groups on these variables, demonstrating the matching was successful (Table 1; Figure 2).

**Figure 2.**
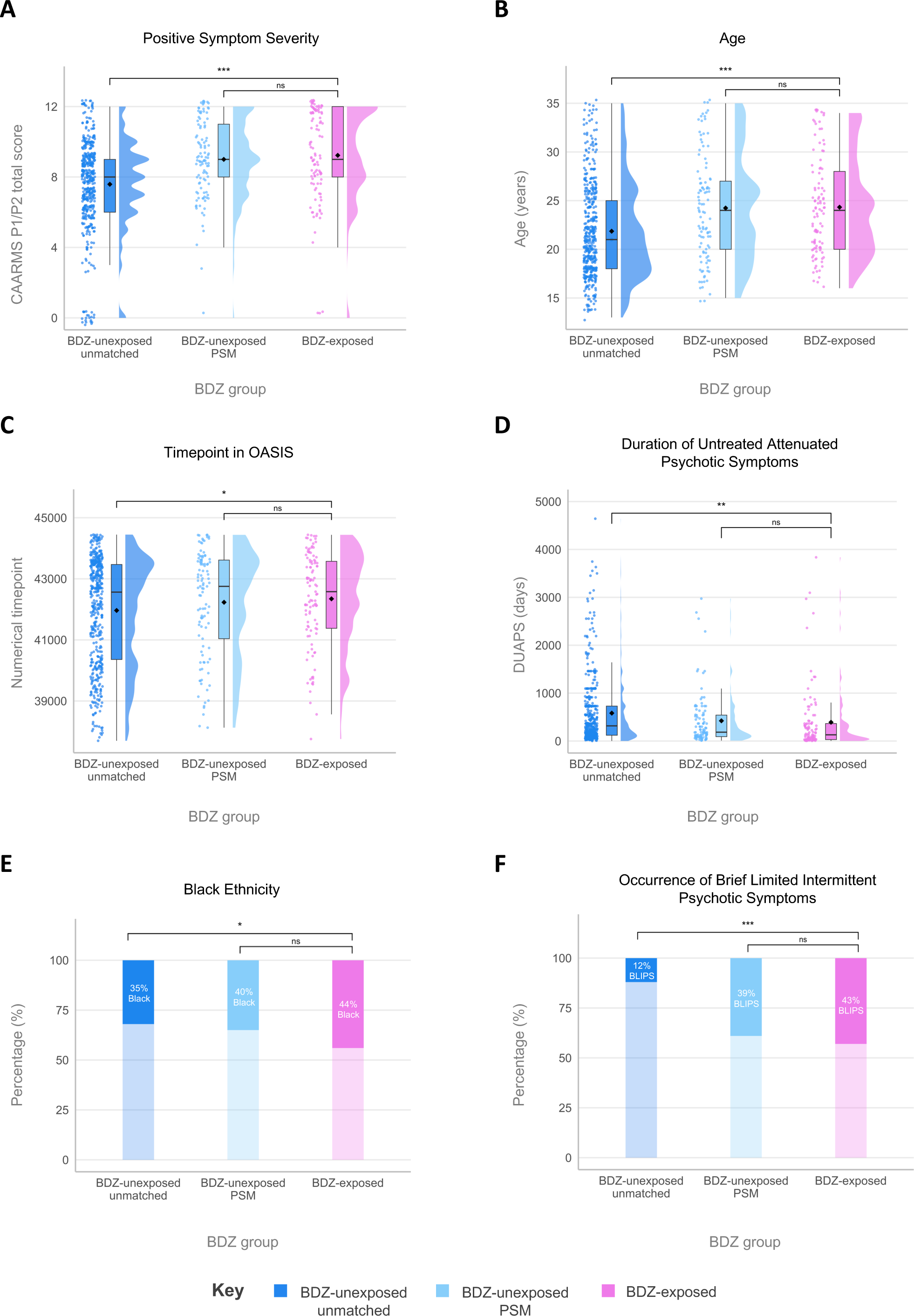
Demographic and Clinical Characteristics Used to Generate Propensity Scores for Propensity Score Matching. Propensity score matching (PSM) successfully matched BDZ-exposed to BDZ-unexposed individuals on characteristics previously associated either with BDZ exposure or psychosis risk (A-F). Dark blue, BDZ-unexposed individuals in the whole sample; light blue, subset generated through PSM; pink, BDZ-exposed individuals. * *P*<.05, ** *P*<.01, *** *P*<.001, ns: non-significant BDZ: Benzodiazepine; BLIPS: Brief Limited Intermittent Psychotic Symptoms; CAARMS: Comprehensive Assessment of At-Risk Mental State; DUAPS: Duration of Untreated Attenuated Psychotic Symptoms; OASIS: Outreach And Support In South London; PSM: Propensity Score Matching

**Table 1.** Demographic and Clinical Characteristics of Individuals in the Whole-Unmatched and Propensity Score Matched Samples.

| Characteristic | BDZ-exposed<br>(n = 105) | BDZ-unexposed<br>unmatched<br>(n = 462) | BDZ-exposed<br>vs. BDZ-unexposed<br>whole-unmatched<br>sample (n = 567) | BDZ-unexposed<br>PSM<br>(n = 105) | BDZ-exposed<br>vs. BDZ-unexposed<br>PSM sample<br>(n = 210) |
| --- | --- | --- | --- | --- | --- |
| | Mean (SD) | Mean (SD) | t/$\chi^2$ (df), *P* | Mean (SD) | t/$\chi^2$ (df), *P* |
| Age (years) | 24.3 (4.9) | 21.9 (4.9) | -4.63 (152.1), <0.001 | 24.2 (5.4) | -0.12 (206.2), 0.91 |
| CAARMS P1-P2 total | 9.2 (2.7) | 7.6 (2.6) | -5.65 (148.3), <0.001 | 9.0 (2.3) | -0.66 (201.2), 0.51 |
| DUAPS (days) | 392 (676) | 639 (891) | 3.16 (195.5), 0.002 | 470 (737) | 0.79 (206.4), 0.43 |
| Timepoint in OASIS | 42345 (1552) | 41964 (1763) | -2.21 (170.6), 0.028 | 42231 (1731) | -0.50 (205.6), 0.62 |
| | Count (%) | Count (%) | t/$\chi^2$ (df), *P* | Count (%) | t/$\chi^2$ (df), *P* |
| **Sex** |  |  | 4.4x10<sup>-31</sup> (1), 1 |  | 0.08 (1), 0.78 |
| Male | 57 (54) | 249 (53.8) | - | 60 (57.1) | - |
| Female | 48 (46) | 213 (46.2) | - | 45 (42.9) | - |
| **Ethnicity** |  |  |  |  |  |
| White | 45 (43) | 213 (46.1) | 0.28 (1), 0.59 | 40 (38.1) | 0.26 (1), 0.61 |
| Asian | 4 (38) | 42 (9.1) | 2.56 (1), 0.11 | 12 (11.4) | 3.39 (1), 0.068 |
| Black<sup>a</sup> | 46 (44) | 146 (31.6) | 5.16 (1), 0.018 | 37 (35.2) | 1.23 (1), 0.26 |
| Other | 10 (10) | 61 (13.2) | 0.59 (1), 0.44 | 16 (15.2) | 0.77 (1), 0.38 |
| **Type of CHR-P subgroup** |  |  |  |  |  |
| APS<sup>b</sup> | 58 (55) | 399 (86) | 21.2 (1), <0.001 | 64 (61) | 0.17 (1), 0.68 |
| GRD | 2 (1.9) | 6 (12.9) | 2.8x10<sup>-4</sup> (1), 0.98 | 0 (0.0) | 0.50 (1), 0.48 |
| BLIPS<sup>c</sup> | 45 (42.9) | 57 (12.3) | 51.97 (1), <0.001 | 41 (39.0) | 0.18 (1), 0.67 |

<sup>a</sup> Black ethnicity includes Black African, Black Caribbean, and Black British
<sup>b</sup> APS also includes APS+GRD
<sup>c</sup> BLIPS also includes BLIPS+APS, BLIPS+GRD and BLIPS+APS+GRD
APS: Attenuated Psychotic Symptoms; BDZ: Benzodiazepine; BLIPS: Brief Limited Intermittent Psychotic Symptoms; CAARMS: Comprehensive Assessment of At-Risk Mental State; CHR-P: Clinical High Risk for Psychosis; DUAPS: Duration of Untreated Attenuated Psychotic Symptoms; GRD: Genetic Risk and Deterioration; OASIS: Outreach And Support In South London; PSM: Propensity Score Matching

### 3.2 Effects of BDZ exposure on clinical outcomes in the whole-unmatched sample

In the whole-unmatched sample (Figure 3A), BDZ exposure was associated with an *increased* risk of transition to psychosis (HR=1.61; 95% CI: 1.03-2.52; *P*=0.037; No. [%] of events for BDZ-exposed vs BDZ-unexposed=26 [24.8%] vs. 75 [16.2%]). Similar results were found for the secondary outcomes, as BDZ exposure was associated with increased risk of psychiatric hospital admission (HR=1.93; 95% CI: 1.13-3.29; *P*=0.017; No. [%] of events BDZ-exposed vs. BDZ-unexposed=19 [17.9%] vs. 45 [9.7%]), home visit (HR=1.64; 95% CI: 1.18-2.28; *P*=0.004; No. [%] of events BDZ-exposed vs. BDZ-unexposed=47 [44.8%] vs. 135 [29.2%]), and A&E attendance (HR=1.88; 95% CI: 1.31-2.72; *P*<0.001; No. [%] of events BDZ-exposed vs. BDZ-unexposed=26 [24.8%] vs. 75 [16.2%]). The proportionality assumption was met for all four models (χ^2^=0.54, *P*=0.46; χ^2^=0.41, *P*=0.52; χ^2^=0.49, *P*=0.48; χ^2^= 15, *P*=0.69, respectively).

**Figure 3.**
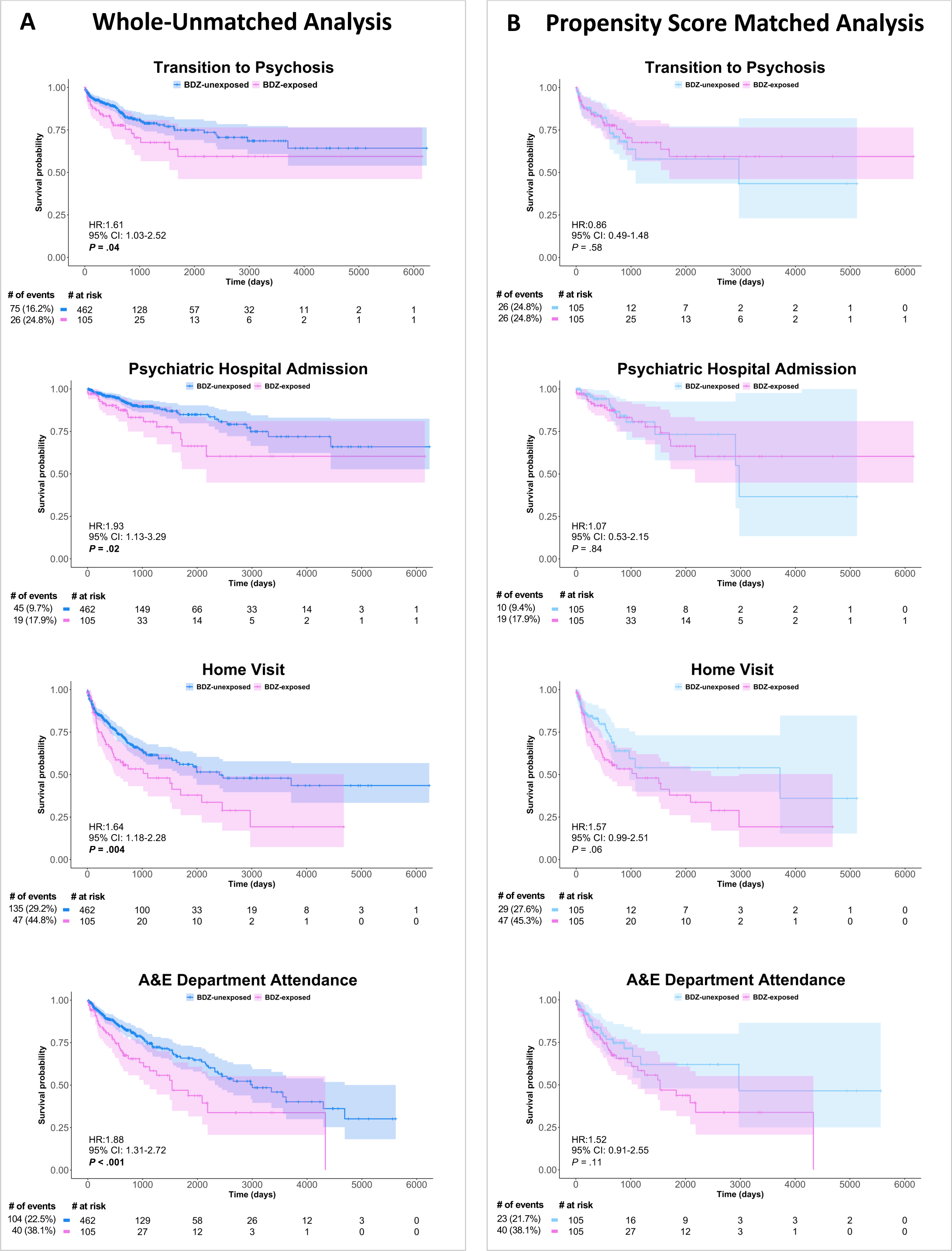
Survival Analysis Comparing the Effect of Benzodiazepine Exposure on the Risk of Subsequent Adverse Clinical Outcomes. Cox proportional-hazards models assessed the effects of BDZ exposure on the four outcome variables for the whole-unmatched sample (A) and PSM sample (B), displayed on Kaplan-Meier survival curves, with the Cox proportional-hazard ratios, confidence intervals and *P* values displayed on each curve and a risk table beneath. N.B. Caution should be used when interpreting Kaplan-Meier curves when <10 individuals are at risk due to high levels of noise. The curves are used for display purposes only, and statistical analyses were only conducted on the cox proportional-hazards models. A&E: Accident and Emergency; BDZ: Benzodiazepine; PSM: Propensity Score Matching

### 3.3 Effects of BDZ exposure on clinical outcomes in the PSM sample

In the PSM sample (Figure 3B), BDZ exposure did not modulate the risk of transition to psychosis (HR=0.86; 95% CI: 0.49-1.48; *P*=0.58; No. [%] of events BDZ-exposed vs. BDZ-unexposed=26 [24.8%] vs. 26 [24.8%]). Similar results were found for the secondary outcomes, as BDZ exposure did not modulate the risk of a psychiatric hospital admission (HR=1.07; 95% CI: 0.53-2.15; *P*=0.84; No. [%] of events BDZ-exposed vs. BDZ-unexposed=19 [17.9%] vs. 10 [9.4%]), home visit (HR=1.57; 95% CI: 0.99-2.51; *P*=0.055; No. [%] of events BDZ-exposed vs. BDZ-unexposed=47 [44.8%] vs. 29 [27.6%]), or A&E attendance (HR=1.52; 95% CI: 0.91-2.55; *P*=0.11; No. [%] of events BDZ-exposed vs. BDZ-unexposed=40 [38.1%] vs. 23 [21.7%]). The proportionality assumption was met for all four models (χ^2^=0.85, *P*=0.36; χ^2^=1.46, *P*=0.23; χ^2^=0.05, *P*=0.83; χ^2^=1.01, *P*=0.31, respectively).

### 3.4 Sensitivity analyses

Removing individuals with prior antipsychotic exposure revealed that BDZ exposure numerically reduced the risk of transition to psychosis, although this did not reach significance (HR=0.59; 95% CI: 0.32-1.08; *P*=0.089; No. [%] of events BDZ-exposed vs. BDZ-unexposed=23 [21.9%] vs. 33 [31.7%]; Figure 4A). BDZ exposure did not modulate the risk of psychiatric hospital admission (HR=0.88; 95% CI: 0.43-1.81; *P*=0.73; No. [%] of events BDZ-exposed vs. BDZ-unexposed=19 [18.3%] vs. 19 [18.3%]; Figure 4B), home visit (HR=1.08; 95% CI: 0.66-1.76; *P*=0.78; No. [%] of events BDZ-exposed vs. BDZ-unexposed=44 [41.5%] vs. 38 [36.6%]; Figure 4C), or A&E attendance (HR=1.73; 95% CI: 0.99-3.02; *P*=0.054; No. [%] of events BDZ-exposed vs. BDZ-unexposed=42 [40.2%] vs. 26 [24.4%]; Figure 4D). The proportionality assumption was met for all four models (χ^2^=1.49, *P*=0.22; χ^2^=3.69, *P*=0.063; χ^2^=1.22, *P*=0.27; χ^2^=1.47, *P*=0.22, respectively). Finally, supplemental sensitivity analyses showed no significant effects of BDZ exposures for 3 or 7 days on any clinical outcomes, and removing non-benzodiazepine hypnotics from the analyses did not change the results, except for a significantly increased risk of A&E attendance (Supplementary Table 2).

**Figure 4.**
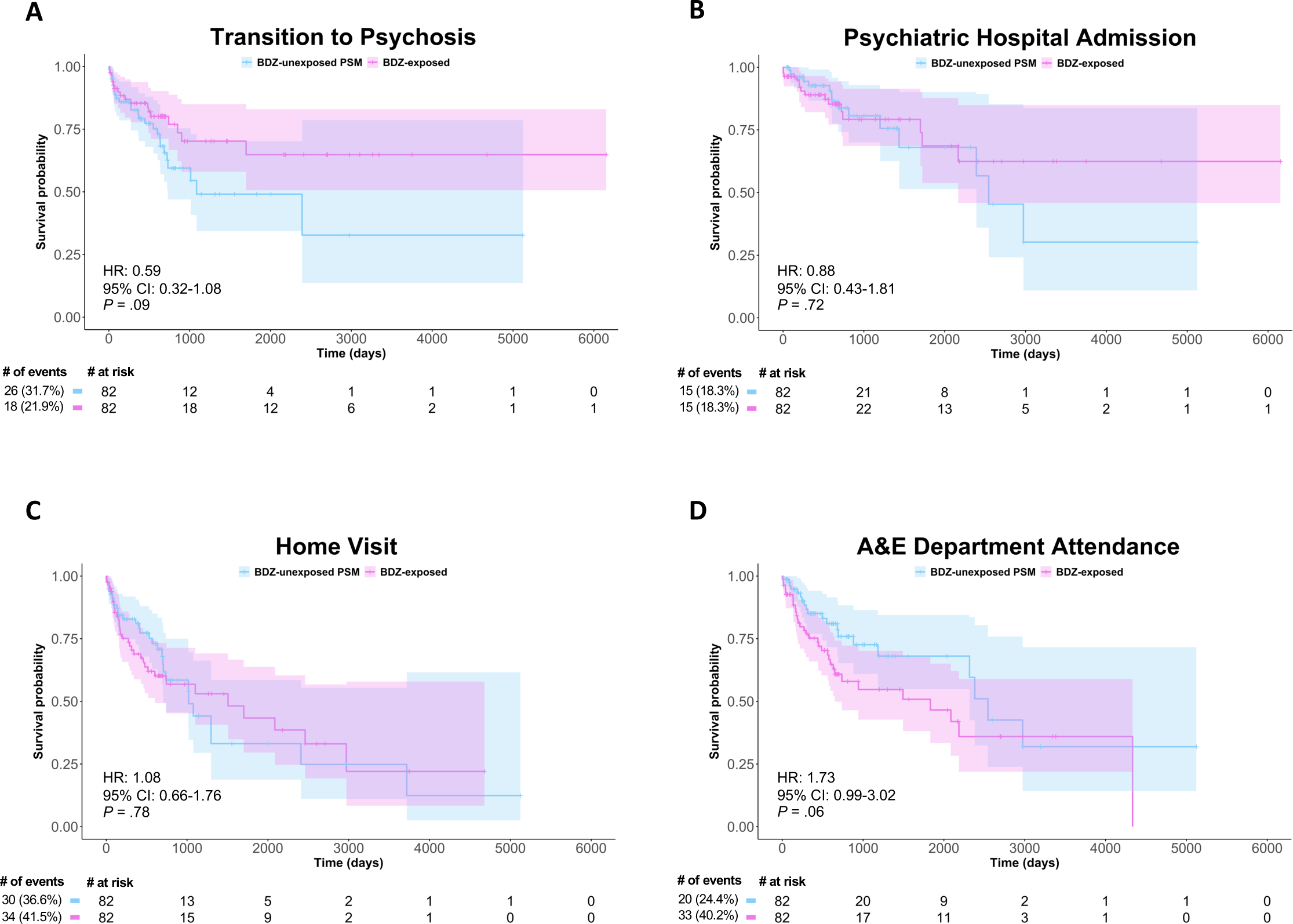
Sensitivity Analysis Removing Individuals with Prior Antipsychotic Exposure. Cox proportional-hazards models assessed the effect of BDZ exposure in antipsychotic-naïve individuals in the PSM sample on the four outcome variables (A-D), displayed on Kaplan-Meier survival curves, with the Cox proportional-hazard ratios, confidence intervals and *P* values displayed on each curve and a risk table beneath. N.B. Caution should be used when interpreting Kaplan-Meier curves when <10 individuals are at risk due to high levels of noise. The curves are used here for display purposes only, and statistical analyses were only conducted on the cox proportional-hazards models. A&E: Accident and Emergency; BDZ: Benzodiazepine; PSM: Propensity Score

## 4. Discussion

To the best of our knowledge, this is the first study using EHR data to investigate the hypothesis that BDZ exposure can improve real-world clinical outcomes in a large sample of CHR-P individuals. In the whole-unmatched sample, BDZ exposure instead increased the risk of developing a psychotic disorder, psychiatric hospital admission, home visit, and A&E attendance. However, after propensity score matching (PSM) to account for confounding-by-indication, BDZ exposure did not modulate the risk of transition to psychosis or other events indicative of a clinical crisis. Restricting the analysis to individuals with no prior antipsychotic exposure suggested that BDZ exposure was associated with a trend-level reduction in the risk of transition to psychosis.

Following PSM, the *increased* risk of transition to psychosis associated with BDZ exposure in the whole-unmatched sample was removed. This suggests confounding-by-indication, such that BDZs are prescribed to individuals who are clinically more unwell, or have a demographic background associated with a higher risk for transition and are therefore already more likely to develop a psychotic disorder. Whilst controlling for these confounds in the PSM analysis removed the increased risk, it did not demonstrate a protective effect of BDZ exposure on psychosis risk as we had hypothesised. Interestingly, when we removed individuals with prior antipsychotic exposure, the hazard ratio for transition to psychosis dropped from 0.86 to 0.59, suggesting a protective effect of the BDZ exposure on psychosis risk in antipsychotic-naïve individuals. We conducted this sensitivity analysis based on preclinical findings that chronic haloperidol treatment blocks the subsequent effects of a selective GABA-enhancing compound (an α5GABA_A_R positive allosteric modulator [PAM]) in MAM-treated rats^23^, likely due to postsynaptic D2-receptor supersensitivity. While this effect in our study was above the threshold for statistical significance, it warrants further investigation due to its potential clinical significance.

The many differences in experimental model and exposure could explain the lack of convergence between preclinical findings and our observations in humans for the primary outcome of transition to psychosis. As with any animal model, the MAM model will have limited validity in capturing the complex neurobiology and outward expressed phenotype of psychosis. Differences in BDZ compound, dose equivalence, and length of dosing period may also be important. We ran several additional sensitivity analyses to increase the equivalence to the preclinical studies, including restricting analyses to individuals with minimum 3 and 7 days of BDZ exposure, and removing BDZ exposures from non-benzodiazepine hypnotics due to the slight differences in pharmacological profiles, but these did not alter the results. The difference in the timing of BDZ exposure is perhaps most relevant to note, as MAM rats were treated peripubertally compared to in young adulthood in our study. For example, environmental enrichment – which has previously demonstrated similar effects to diazepam in preventing the psychosis phenotype in the MAM model – is not effective when given postpubertally compared to peripubertally^33^. There are also much higher levels of heterogeneity between CHR-P individuals than in MAM rats. Only approximately 25% of CHR-P individuals will transition to psychosis^6^, and the CHR-P state is associated with a multitude of potential trajectories in terms of symptoms and functioning^8^ which may be driven by differences in pathophysiology. Preclinical studies have demonstrated that the pharmacological effect of GABAergic treatment on the subcortical dopaminergic system differs between control and MAM rats^23^, and correspondingly BDZ effects might differ between CHR-P individuals. The heterogeneity between CHR-P individuals may also explain why no other effective preventative treatment has been discovered^14,34^.

Investigation of secondary outcomes after PSM also found no positive effects of BDZ exposure on the risk of real-world events indicative of a clinical crisis in CHR-P individuals, including psychiatric hospital admission, home visit, and A&E attendance. In fact, findings indicated a residual increased risk of receiving a home visit following BDZ exposure, albeit non-significant. Additionally, in our sensitivity analyses, there was a trending increased risk of subsequent A&E attendance by BDZ exposure when analysis was restricted to antipsychotic-naïve individuals, and a significant increased risk on A&E attendance when BDZ exposures from a non-benzodiazepine hypnotic were removed from analysis. Whilst this is the first study to examine the influence of BDZ exposure on clinical outcomes in CHR-P individuals, a recent EHR study in first-episode psychosis patients investigated the effects of antipsychotic and BDZ treatment (within the first week of illness onset) on clinical outcomes and found similar effects^35^. BDZ treatment prior to antipsychotic treatment (vs. after) increased the duration of medical and A&E admission in first-episode patients, whilst reducing the length of psychiatric admission^35^. Furthermore, increased readmission to hospital has been associated with BDZ exposure in patients with chronic schizophrenia^36,37^. An increased risk of home visit, A&E attendance, or hospitalisation might reflect residual confounding-by-indication as these events capture non-specific clinical crises influenced by a multitude of factors. For example, comorbid anxiety disorder may be associated with both BDZ exposure and repeated presentations at A&E with consequential hospital admission, creating a false association between the exposure and the event. Differences in the reason for prescription of non-benzodiazepine hypnotics compared to traditional benzodiazepines (sleep difficulties and anxiety/agitation, respectively) might explain why removing non-benzodiazepine hypnotics exposed individuals from the analysis led to a significant increased risk of A&E attendance from BDZ exposure, as the analysis was restricted to individuals with a clinical profile more likely to present to A&E. Alternatively, these worse clinical outcomes could be influenced by adverse effects of BDZs including psychotic features and other adverse behavioural effects^38^, which is important to note as in this study we do not know whether A&E attendances were related to psychiatric or other medical events. However as these effects are very rare (< 1%)^39^, they are unlikely to be driving these findings in our sample.

This study has several strengths. We investigated the effects of BDZ exposure on transition to psychosis and clinical outcomes in CHR-P individuals, informing clinical understanding of effectiveness and safety of BDZ in this population. Secondly, we used real-world data with high ecological validity from one of the longest established CHR-P services, affording a large sample size from a residential population with one of the highest psychosis rates worldwide^40^. Thirdly, we used advanced statistical methods including PSM to account for confounding-by-indication, and in the process characterised disparities in BDZ exposure between CHR-P individuals which were not previously established.

This study also has some limitations. Firstly, despite using PSM to control for confounding factors, observational studies with real-world data are susceptible to confounding-by-indication. We controlled for several factors associated with BDZ exposure and adverse clinical outcomes as identified in prior research, but it is likely that there is residual confounding from additional factors that we were not statistically powered to include (e.g., cannabis use). Secondly, the observational nature of the study means that factors such as treatment compliance are not known, which could impact clinical outcomes. Additionally, subsequent clinical care (e.g., pharmacological/psychological interventions) beyond the exposure window but before the occurrence of events was not measured. Finally, limitations of the translatability of preclinical findings outlined earlier in terms of differences in BDZ compound, dose equivalence, dosing period, and timing of BDZ exposure are relevant to note.

## Conclusions

In conclusion, BDZ exposure in CHR-P individuals was not associated with a reduced risk of developing psychosis or adverse clinical outcomes after controlling for confounding-by-indication. We found suggestive evidence that prior antipsychotic exposure could be attenuating the potential therapeutic effects of BDZs in this clinical population. Further experimental research in this field is warranted, to investigate the effect of post-pubertal BDZ administration in preclinical models, investigate real-world data from child and adolescent mental health services to capture an even earlier developmental time window, and develop more selective GABAergic agents (e.g., α5GABA_A_R PAM) with better side-effect profiles and which more specifically target areas of neurobiological dysfunction in CHR-P individuals such as the hippocampus.

## Data Availability

There is no ethical permission for data or research material sharing. Code is available upon request.

## Acknowledgments

NRL is supported by a PhD studentship from the Medical Research Council Doctoral Training Partnership. This research was funded in whole, or in part, by the Wellcome Trust & The Royal Society [Sir Henry Dale Fellowship 202397/Z/16/Z to GM]. RAMs work is funded by a Wellcome Trust Clinical Research Career Development (224625/Z/21/Z). AAG’s work is funded by a United States Public Health Service Grant (NIMH MH57440). For the purpose of open access, the author has applied a CC-BY public copyright licence to any Author Accepted Manuscript version arising from this submission.

