## Supplementary Material for "Effects of Benzodiazepine Exposure on Real-World Clinical Outcomes in Individuals at Clinical High-Risk for Psychosis"

**Supplementary Table 1. Details of Benzodiazepine Exposures Included in Analysis (n = 105)**

**Supplementary Table 2. Additional Sensitivity Analyses in the Propensity Score Matched Sample**

**Supplementary Table 1. Details of Benzodiazepine Exposures Included in Analysis (n = 105)**

| | | **Count (%)** |
|---|---|---|
| **BDZ name** | | |
| | Clonazepam | 16 (15.1) |
| | Lorazepam | 12 (11.3) |
| | Diazepam | 16 (15.1) |
| | Temazepam | 1 (0.9) |
| | Zopiclone/Zolpidem | 41 (38.7) |
| | Alprazolam | 1 (0.9) |
| | Bromazepam | 1 (0.9) |
| | Clonazepam + Lorazepam | 1 (0.9) |
| | Clonazepam + Diazepam | 3 (2.8) |
| | Clonazepam + Zopiclone | 7 (6.6) |
| | Lorazepam + Diazepam | 1 (0.9) |
| | Lorazepam + Zopiclone | 1 (0.9) |
| | Diazepam + Zopiclone | 4 (3.8) |
| | Bromazepam + Zopiclone | 1 (0.9) |
| **Reason for BDZ exposure** | | |
| | Anxiety | 17 (16.0) |
| | Sedation | 3 (2.8) |
| | Agitation | 13 (12.3) |
| | Sleep | 59 (55.7) |
| | Not known | 13 (12.3) |
| | | **Mean (± SD)** |
| **Total number of BDZ exposure (days)** | | 18.5 (25.6) |

BDZ: Benzodiazepine

**Supplementary Table2. Additional Sensitivity Analyses in the Propensity Score Matched Sample**

| | | Transition to Psychosis | Psychiatric Hospital Admission | Home Visit | A&E Attendance |
|---|---|---|---|---|---|
| **≥ 3 total days of BDZ (n = 89 per group)** | | | | | |
| | HR (95% CI), *P* | 1.67 (0.89-3.11), .11 | 1.67 (0.79-3.53), .18 | 1.38 (0.86-2.23), .18 | 1.23 (0.75-2.18), .37 |
| | Number of events; BDZ exposed vs. unexposed | 22 vs. 18 | 16 vs. 12 | 37 vs. 31 | 33 vs. 22 |
| **≥ 7 total days of BDZ (n = 66 per group)** | | | | | |
| | HR (95% CI), *P* | 1.61 (0.75-3.45), .22 | 0.82 (0.32-2.12), .69 | 1.65 (0.94-2.89), .08 | 1.11 (0.56-2.19), .77 |
| | Number of events; BDZ exposed vs. unexposed | 11 vs. 7 | 7 vs. 4 | 24 vs. 12 | 16 vs. 10 |
| **Removing non-benzodiazepine hypnotics (n = 75 per group)** | | | | | |
| | HR (95% CI), *P* | 0.91 (0.47-1.77), .79 | 1.41 (0.58-3.41), .44 | 1.47 (0.87-2.49), .15 | 2.08 (1.11-3.93), **.02** |
| | Number of events; BDZ exposed vs. unexposed | 18 vs. 17 | 13 vs. 8 | 41 vs. 28 | 31 vs. 16 |

A&E: Accident & Emergency; BDZ: Benzodiazepine
